# Low somatosensory cortex excitability in the acute stage of low back pain causes chronic pain

**DOI:** 10.1101/2021.02.18.21251719

**Authors:** Luke C Jenkins, Wei-Ju Chang, Valentina Buscemi, Matthew Liston, Patrick Skippen, Aidan G Cashin, James H McAuley, Siobhan M Schabrun

## Abstract

**BACKGROUND:** Determining the mechanistic causes of complex biopsychosocial health conditions such as low back pain (LBP) is challenging, and research is scarce. Cross-sectional studies demonstrate altered excitability and organisation of the primary somatosensory and primary motor cortex in people with acute and chronic LBP, however, no study has explored these mechanisms longitudinally or attempted to draw causal inferences.

**METHODS:** Prospective, longitudinal, cohort study including 120 people with an acute episode of LBP. Sensory evoked potential area measurements were used to assess primary and secondary somatosensory cortex excitability. Transcranial magnetic stimulation derived map volume was used to assess corticomotor excitability. Directed acyclic graphs identified variables potentially confounding the exposure-outcome relationship. The effect of acute-stage sensorimotor cortex excitability on six-month LBP outcome was estimated using multivariable regression modelling, with adjusted and unadjusted estimates reported. Sensitivity analyses were performed to explore the effect of unmeasured confounding and missing data.

**RESULTS:** Lower primary (OR = 2.08, 95% CI = 1.22 to 3.57) and secondary (OR = 2.56, 95% CI = 1.37 to 4.76) somatosensory cortex excitability in the acute stage of LBP increased the odds of developing chronic pain at six-month follow-up. This finding was robust to confounder adjustment and unmeasured confounding (E-Value = 2.24 & 2.58, respectively). Corticomotor excitability in the acute stage of LBP was associated with higher pain intensity at 6-month follow-up (B = −0.15, 95% CI: −0.28 to −0.02) but this association did not remain after confounder adjustment.

**CONCLUSION:** These data provide the first evidence that low somatosensory cortex excitability in the acute stage of LBP is a cause of chronic pain. Interventions designed to increase somatosensory cortex excitability in acute LBP may be relevant to the prevention of chronic pain.

## 1. INTRODUCTION

Low back pain (LBP) is the leading cause of years lost to disability worldwide (1) with an economic burden comparable to cardiovascular disease, cancer, mental health, and autoimmune diseases (2). Up to 40% of people who experience an episode of acute LBP develop chronic symptoms (3) and interventions designed to prevent the development of chronic LBP have not been effective (4-8). A critical issue is the lack of robust mechanistic explanations for chronic LBP; in 85 – 95% of LBP cases (9-11), a pathoanatomical cause cannot be determined and the condition is labelled ‘non-specific’ (12, 13). Identifying the causal mechanisms of chronic LBP is a recognised research priority that would guide the development of targeted treatment and prevention strategies.

Determining the mechanistic causes of complex biopsychosocial health conditions such as LBP is challenging, and research is scarce. Psychological factors, such as distress, pain-related fear and low self-efficacy, have been identified as potential causes of persistent disability following acute LBP (14). However, studies investigating these factors frequently use cross-sectional designs and rarely control for confounding variables, leading some experts to suggest these factors are a consequence, rather than a cause, of chronic LBP (14, 15). Interventions attempting to prevent the development of chronic LBP through reduction of psychological risk factors have also been ineffective (4, 6). Taken together, these data suggest other mechanisms must play a role in the development of chronic LBP (4, 14).

Aberrant sensorimotor cortex excitability in the acute stage of LBP is one mechanism hypothesised to have a causal relationship with chronic LBP (16-18). Cross-sectional studies demonstrate altered excitability and organisation of the primary somatosensory (S1) and primary motor (M1) cortex in people with acute and chronic LBP (19, 20) and these changes are associated with pain severity, functional impairment and symptom chronicity (21-24). For example, processing of non-noxious sensory inputs is supressed, and corticomotor excitability lower, in people with acute LBP compared with healthy controls, and individuals who display lower S1 excitability in the acute stage of LBP experience worse pain than those who display higher S1 excitability (25). Further, longitudinal research using experimental pain models suggests that individuals who display low corticomotor excitability soon after pain onset experience worse pain and slower recovery than those who display high corticomotor excitability (26). Despite these findings, no study has investigated whether there is evidence for a causal relationship between acute-stage sensorimotor cortex excitability and the development of chronic LBP (27).

Recent conceptual advances have outlined methods for estimating the causal effect of an exposure on a health outcome using observational data (28-30). Using longitudinal data obtained from the UPWaRD (Understanding persistent Pain Where it ResiDes) cohort, we implemented these conceptual advances to investigate the causal relationship between acute stage sensorimotor cortex excitability and LBP outcome at six-month follow-up. Directed acyclic graphs (DAGs) were used to identify possible confounders of the exposure (sensorimotor cortex excitability) -outcome (pain and disability at six months) relationship and minimise confounding bias. We hypothesised that acute-stage sensorimotor cortex excitability would demonstrate a causal relationship with chronic LBP when confounders were controlled.

## 2. METHODS

### 2.1. Study design

The UPWaRD study was a multicentre, prospective, longitudinal, cohort study of people presenting with acute LBP (National Health and Medical Research Council of Australia, Grant ID: 1059116). Participants underwent a battery of neurophysiological and psychological tests at baseline (within 6 weeks of pain onset) with follow-up at six-months. Consistent with recommendations for transparency and reproducibility (31, 32) the protocol for data collection and the statistical analysis plan were registered with the Australian and New Zealand Clinical Trials Registry (ACTRN12619000002189) and published *‘a priori’* (33, 34). This study reports the findings from the analysis plan in accordance with the STROBE guidelines (35). Any deviation from the planned analyses is noted.

### 2.2. Study Population

One hundred and twenty people experiencing an acute episode of LBP were recruited from local hospitals in South Eastern and South Western Sydney local health districts, New South Wales, Australia, primary care practitioners (e.g. general practitioners and physiotherapists), newspaper/online advertisements, flyers and social media sites such as Facebook. Participants were included if they experienced pain in the region of the lower back, superiorly bound by the thoracolumbar junction and inferiorly by the gluteal fold (36). Participants remained eligible for inclusion if they had pain referred beyond this region that was not radicular pain from neural tissue involvement. Pain must have been present for more than 24 hours and persisted for less than six weeks following a period of at least one-month pain-free (36-39). All participants with pain referred beyond the inferior gluteal fold underwent a physical examination by a trained physiotherapist (study staff) to identify any sensory or motor deficit of the lower extremity. Participants with suspected lumbosacral radiculopathy characterised by the presence of weakness, loss of sensation, or loss of reflexes associated with a particular nerve root, or a combination of these, were excluded (40). Individuals who presented with suspected serious spine pathology (e.g. fracture, tumour, cauda equina syndrome), other major diseases/disorders (e.g. schizophrenia, chronic renal disorder, multiple sclerosis), a history of spine surgery, any other chronic pain conditions or contraindications to the use of transcranial magnetic stimulation (TMS) were excluded (41). Four assessors performed all study related procedures at laboratories located at Western Sydney University or Neuroscience Research Australia, New South Wales, Australia. All procedures were approved by Western Sydney University (H10465) and Neuroscience Research Australia (SSA: 16/002) Human Research Ethics Committees and conducted in accordance with the Declaration of the World Medical Association (42). All patients gave written informed consent.

### 2.3. Primary and secondary outcomes

Pain and disability were assessed six months after the baseline assessment to determine if participants had developed chronic LBP. The primary outcome was pain intensity and the secondary outcome disability (33). Participants completed the Brief Pain Inventory (43) and were asked to score their pain intensity on average over the previous week using an 11-point numerical rating scale (numeric rating scale [NRS]: 0=‘no pain’, 10=‘worst pain imaginable’). Participants were considered to have developed chronic pain if they reported a NRS score ≥ 1 at six-month follow-up (44). Disability was assessed using the 24-Item Roland Morris Questionnaire (RMDQ) scored from 0 (no disability) to 24 (high disability) (45). Participants who scored ≥ 3 on the RMDQ at six-month follow-up were considered to have developed chronic disability (44). We chose these stringent definitions as we were interested in ‘true’ recovery i.e. individuals who reported no on-going pain or disability at 6-months follow-up.

### 2.4. Exposure variables: Sensorimotor cortex excitability

Sensory evoked potentials (SEPs) were recorded using gold plated cup electrodes positioned over S1 (3cm lateral and 2cm posterior of Cz) on the hemisphere contralateral to the side of worst LBP and referenced to Fz using the International 10/20 System (46). The side of worst LBP was determined on the day of baseline testing by asking the participant “on average over the past 24 hours which side of your back is most painful?” If the participant was unable to determine a more painful side, and reported central LBP at all times over the previous 24 hours, SEPs were recorded on the hemisphere contralateral to the dominant hand (16, 47). Electrode impedance was maintained at <5 k*Ω*. Electroencephalography (EEG) signals were amplified 50,000 times, band pass filtered between 5 and 500Hz, and sampled at 1,000 Hz using a Micro1401 data acquisition system and Signal software (CED Limited, Cambridge, UK).

SEPs were recorded in response to two blocks of 500 non-noxious electrical stimuli. Participants were seated comfortably in a chair with feet on the floor and arms relaxed. A single bipolar electrode (silver-silver chloride disposable electrodes; Noraxon USA, Arizona, USA) was positioned 3cm lateral to the L3 spinous process, ipsilateral to the side of the worst LBP and a constant current stimulator (Digitimer Ltd, Hertfordshire, UK DS7AH) delivered the electrical stimulation. Anodal stimulation was applied to the inferior attachment of the bipolar electrode. Stimulation was increased in 1mA increments until the perceptual threshold was reached. The testing intensity was set at three times perceptual threshold. If this intensity evoked pain, it was decreased in 1mA increments until the stimulus became non-noxious. The electrical stimuli had a pulse duration of 1ms and were delivered at a frequency of 2Hz with a variable interval schedule of 20%. Participants were asked to sit still with their eyes closed, but remain awake, during the procedure.

Individual SEP traces were manually inspected and those considered to contain eye movements, muscle artefacts or electrical interference were rejected. Less than 15% of all SEP traces were excluded. Remaining traces from the two SEP blocks were averaged for each participant and the average used for analysis (16). The averaged wave form was full-wave rectified and the area under the curve mean amplitude (µV) determined for the N_80_ (between the first major downward deflection of the curve after stimulus onset and the first peak, N_80_) and N_150_(between the first and second peak, N_80_ and N_150_ respectively) time windows. The SEP area measurement was chosen for analysis as it is less susceptible to signal-to-noise ratio concerns (48), and considered superior to peak-based measures for assessing event-related potentials (17, 49-51). Previous electrophysiological research suggests distinct EEG components reflect sensory afferent processing within discrete cortical regions (52) -the N_80_ SEP time window is thought to reflect processing in S1, while the N_150_ SEP time window is thought to reflect processing in the secondary somatosensory cortex (S2) (17, 21).

The corticomotor response to TMS was assessed using a standardised mapping procedure (53-55). Surface electromyography (EMG) was recorded from the paraspinal muscles 3 cm lateral to the spinous process of L3 and 1 cm lateral to the spinous process of L5 using disposable Ag/AgCL electrodes (Noraxon USA Inc, Arizona, USA) (56, 57). Ground electrodes were placed over the anterior superior iliac spine bilaterally. EMG data were amplified 1000x, filtered 20-1000 Hz and sampled at 2000 Hz using a Micro1401 data acquisition system and Spike2 software (CED Limited, Cambridge, UK).

Single-pulse, monophasic stimuli (Magstim 200 stimulator/7 cm figure-of-eight coil; Magstim Co. Ltd. Dyfed, UK) were delivered over M1 contralateral to the side of the worst LBP. The coil was placed tangentially to the skull with the handle pointing posterior-laterally at 45 degrees from midline (58-60). Participants wore a cap marked with a 6 x 7 cm grid oriented to the vertex (point 0, 0). The vertex was determined using the International 10/20 System, and aligned with the centre of the cap (61). The cap was tightly fitted and the position regularly checked to ensure placement consistency. Starting at the vertex, five stimuli were delivered over each site on the grid with an inter-stimulus interval of 6 s. As 120% of active motor threshold for paraspinal muscles often exceeds the maximum stimulator output, all stimuli were delivered at 100% while participants activated the paraspinal extensor muscles to 20 ± 5% of their EMG recorded during a maximum voluntary contraction (MVC) (determined as 20% of the highest root mean square [RMS] EMG for 1 s during three, 3 s maximal muscle contractions performed against manual resistance in sitting) (54, 62, 63). Feedback of real-time RMS EMG of paraspinal extensor muscles and the target level was displayed on a monitor (64). All TMS procedures adhered to the TMS checklist for methodological quality (65).

TMS map data were analysed offline using MATLAB 2019a (The MathWorks, USA). Motor evoked potential (MEP) onset and offset for each individual trace were visually identified then averaged at each scalp site. The amplitude of paraspinal MEPs was measured as the RMS EMG amplitude, between the onset and offset of the MEP, from which the background RMS EMG was removed (55–5 ms preceding stimulation) (54). Paraspinal MEP amplitudes were normalized to the peak MEP amplitude and superimposed over the respective scalp sites to generate a topographical map. A scalp site was considered active if the normalised MEP amplitude was equal to or greater than 25% of the peak response (16). Normalised values below 25% of the peak response were removed and the remaining values rescaled from 0 to 100% (53, 66). Map volume was calculated as the sum of normalized MEP amplitudes recorded at all active scalp sites (67).

### 2.5. Identifying sources of confounding

Variables thought to confound the relationship between an acute episode of LBP and the development of chronic LBP were graphically represented using causal directed acyclic graphs (DAGs). DAGs incorporate expert content knowledge to explicitly represent assumptions about the causal relationships between variables, providing a non-parametric framework to identify the minimum sufficient set of variables that must be measured and controlled to obtain unconfounded causal effect estimates (68). Following a meeting between study investigators and content experts, causal DAGs were developed using DAGitty (69) and the following variables were identified as confounders: predisposing factors, physical activity, baseline symptoms, comorbidity, sensitisation, blood biomarkers, treatment, hypothalamic-pituitary-adrenal axis/cortisol and psychological variables (**Figure 1**). The sufficient set of covariates to minimise confounding bias included predisposing factors, blood biomarkers, psychological variables and sensitisation. Once variables identified as the sufficient set from the DAG were controlled, no alternate paths between the exposure and the outcome remained open (28), (**Figure 2**).

**Figure 1.**
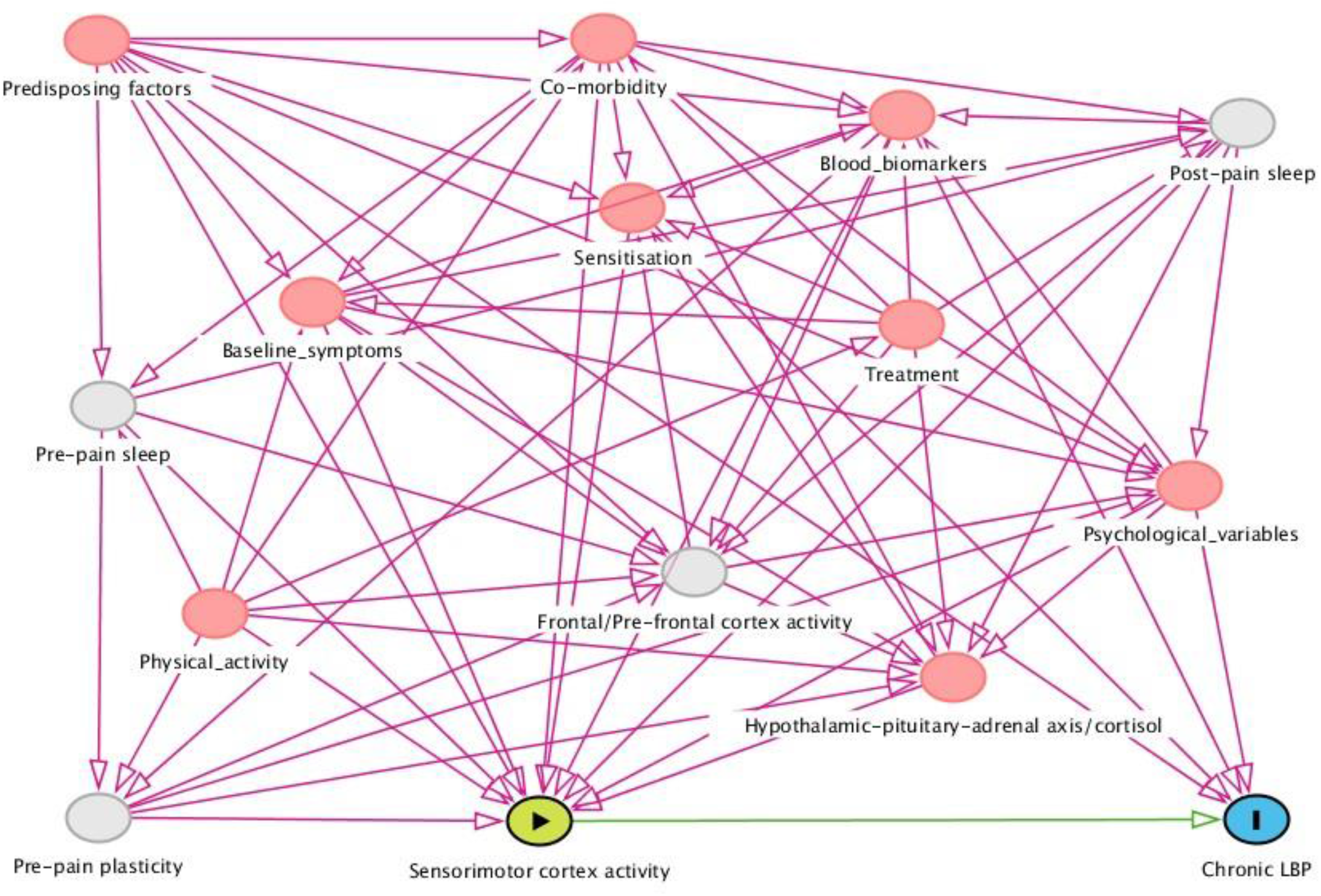
Directed Acyclic Graph to identify confounding variables. Confounding variables are in red boxes. Blue box is the outcome. Green box is the exposure. Clear circles are variables that were unobserved yet assumed to have a causal effect on exposure and outcome.

**Figure 2.**
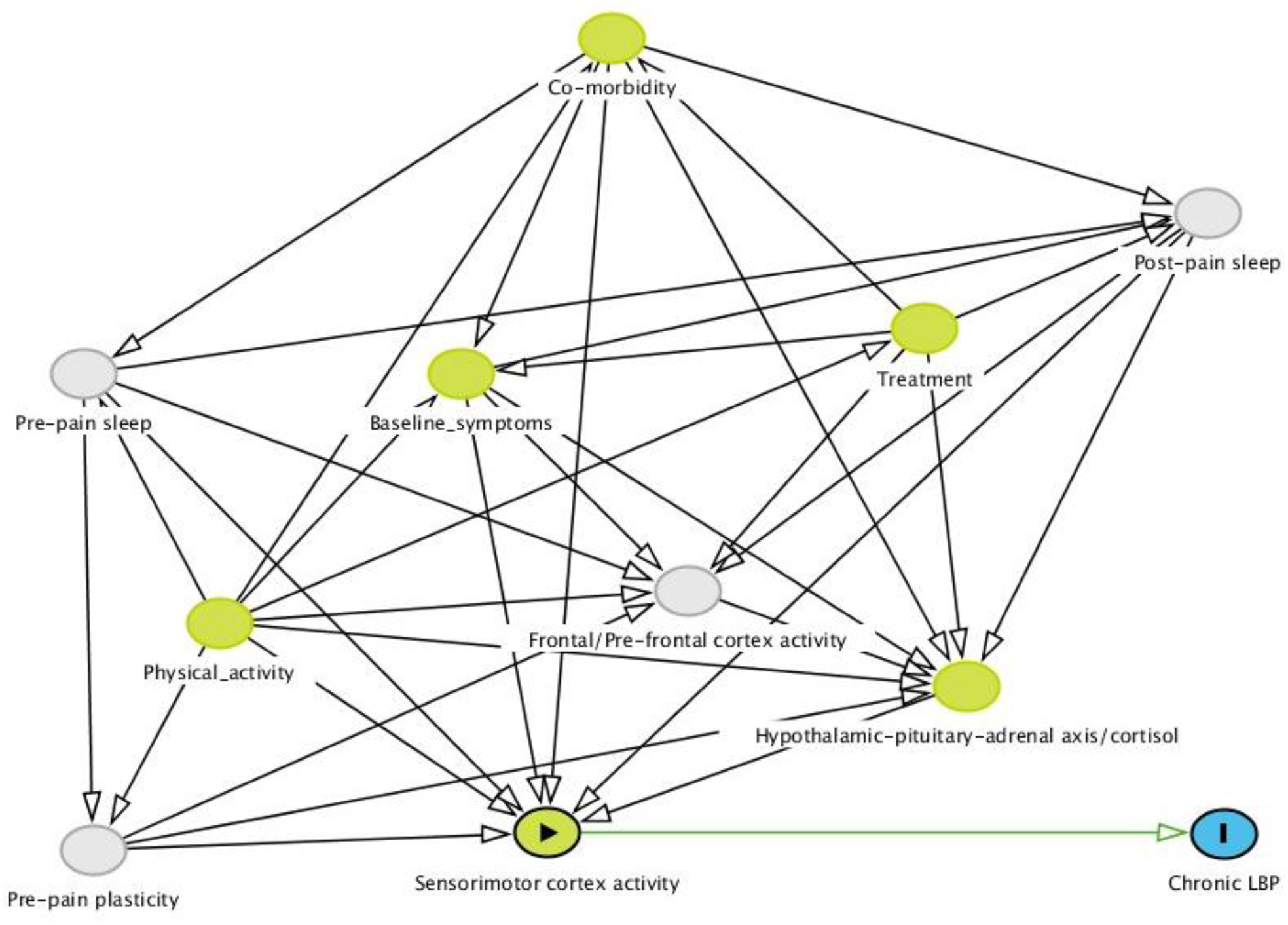
Directed Acyclic Graph confirming there are no ‘back door’ causal paths in the DAG. Variables identified in Figure 1 as the ‘sufficient’ set have been removed. Blue box is the outcome. Green boxes are variables that only have a causal effect on the exposure. Clear circles are variables that were unobserved yet assumed to have a causal effect on exposure and outcome.

#### 2.5.1. Predisposing factors

A range of predisposing factors were assessed including age, sex, previous history of LBP, socioeconomic status and cultural and linguistic diversity. Further, heritability of LBP is estimated to be between 30 – 60% (70, 71), therefore, brain-derived neurotrophic factor (BDNF), a high priority candidate gene thought to encode numerous mediators of pain processing was also considered a predisposing factor (72-74). Participants were considered to have a previous history of LBP if they answered “yes” to the question: “Have you experienced low back pain in the past”? Each participant’s postal code was converted into a socioeconomic index for areas (SEIFA) score, with higher scores representing, on average, higher socioeconomic status for people living within that postal code (75). Each participant was asked the question: “How do you define your identity, in ethnic or cultural terms?” If the participant identified a cultural or ethnic background other than “English”, “Caucasian” or “Australian” they were considered culturally and linguistically diverse for the purpose of this study. To determine each participants BDNF genotype, buccal swabs were taken on the day of baseline testing (Isohelix DNA Isolation Kit) and used to prepare genomic DNA samples (76). Samples were immediately frozen and stored at −80°C. Samples were polymerase chain reaction-amplified and sequenced by the Australian Genome Research Facility (AGRF); see (77). Consistent with prior investigations (73, 78, 79), BDNF was coded as a dichotomous variable (AA/AG or GG). The more common G allele encodes the Val, while the A allele encodes Met. In the current sample, 72 (60%) participants were coded GG and 48 (40%) coded AA/AG. According to the Hardy–Weinberg Equilibrium, this observed distribution is consistent with the expected rate (χ^2^ = 1.27, df = 1, *P* = 0.26).

#### 2.5.2. Blood biomarkers

Peripheral venous blood was drawn into serum tubes (BD, SST II Advance) through venepuncture of the median cubital vein by a phlebotomy-trained member of the research team at baseline assessment. The sample was clotted (30 min, room temperature) then separated by centrifugation (2500 rpm, 15 min). The samples were pipetted into 50 μL aliquots and stored at −80°C until analysis. After thawing, the Simple plex Ella™ platform was used to analyse the specific expression of C-reactive protein (CRP), tumour necrosis factor (TNF), interleukin-1β (IL-1β), interleukin-6 (IL-6) and circulating BDNF. Samples were prepared and loaded into the cartridge according to a standard procedure provided by the manufacturers (Protein Simple, CA, USA). All steps in the immunoassay procedure were carried out automatically and scans were processed with no user activity. Cartridges included built-in lot–specific standard curves. Single data (pg/mL) for each sample were automatically calculated. The limits of detection for each biomarker were as follows: (1) CRP: 1.24 pg/ml, (2) TNF-α: 0.278 pg/ml, (3) IL-1β: 0.064 pg/ml, (4) IL-6: 0.26 pg/ml and (5) BDNF: 5.25 pg/ml. Zero was allocated for values below the test sensitivity.

#### 2.5..3. Psychological variables

Psychological screening questionnaires were administered to each participant at baseline assessment and included the total score of the 21-item depression, anxiety, stress subscale (DASS-21) (80), pain catastrophizing scale (PCS) (81), and pain self-efficacy questionnaire (PSEQ) (82). These self-reported questionnaires measure emotional and cognitive domains of the participant’s pain experience.

#### 2.5.4. Sensitisation

Pressure pain thresholds (PPTs) were measured locally (3cm lateral to the L3 spinous process, ipsilateral to side of greatest LBP) and distally (thumbnail of the hand ipsilateral to side of greatest LBP) to represent peripheral and widespread mechanical pain sensitivity, respectively. A hand-held pressure algometer with probe size 1cm^2^ (Somedic, Hörby, Sweden), was applied perpendicular to the skin and the participant reported when the sensation first changed from pressure to pain. The average of three trials was used for analysis.

### 2.6. Statistical analyses

G*Power (V.3.0.10, University of Kiel, Germany) was used to calculate the required sample size for estimating the causal effect of acute stage sensorimotor cortex excitability on outcome (83). According to the sample size calculation, 111 participants were required to detect an effect size of 0.2 with 80% power, using an alpha level of 0.05, with 16 confounding variables included within the sufficient adjustment set. This calculation is based on detecting a medium effect for a multiple linear regression (84).

Statistical analyses were performed using R software (The R Foundation for Statistical Computing, a statistical software) (85). All missing data were imputed using the Multivariate Imputation by Chained Equations (MICE) procedure (52). The imputation model was adapted to the type of variables. Incomplete dichotomous variables were imputed using a logistic regression model, while predictive mean matching was used to impute incomplete continuous variables. All variables displayed normal distribution except the N_80_ and N_150_ SEP area and serum concentration of CRP. Therefore, these variables were first log-transformed before conducting further analyses. All available data, including the outcome variables were used in the imputation procedure (86). Thirty imputed data sets were generated. All analyses were performed on each imputed dataset and results were pooled using Rubin’s rules (87).

Baseline data for both the exposure and confounding variables were compared between those who developed chronic pain or chronic disability and those who recovered at follow-up using chi-squared (categorical variables) or independent t tests (continuous variables). Homogeneity of variance was assessed using Levene’s test and for variables that did not meet the equal variance assumption a Welch’s t test was performed.

To investigate the effect of acute-stage sensorimotor cortex excitability on pain intensity and disability, linear regression models were created. In these models, pain intensity or RMDQ score was entered as the continuous dependent variable of interest and N_80_ area, N_150_ area, L3 map volume or L5 map volume was entered as the independent variable. Unstandardized beta coefficients were reported for the linear models. Logistic regression models were used to investigate the effect of sensorimotor cortex excitability on chronic pain or chronic disability. The dichotomized NRS or RMDQ score was entered as the dependent variable and odds ratios (OR) reported. Confounders identified by the DAG were entered into the models as covariates and adjusted for in the final estimate of effect. False Discovery Rate (FDR) corrections were performed on all *P*-values within each model. This correction controls for the expected fraction of significant tests (i.e., *P* < 0.05) in which the null hypothesis would actually be true (88).

A sensitivity analysis was performed to explore the influence of unmeasured confounding on the observed causal effect (89). This was achieved by calculating the E-value, using the R package: “EValue” (89). The E-value defines the minimum strength of association that an unmeasured confounder would need to have with both sensorimotor cortex evoked excitability and chronic LBP to fully explain away the exposure-outcome relationship, conditional on measured covariates (89). A secondary sensitivity analysis was completed to explore the effect of missing data. In this analysis (see Appendix 1) the statistical analysis plan was repeated on complete cases, without imputation.

## 3. RESULTS

### 3.1. Study flow

Between April 2015 and January 2019, 498 participants who presented with LBP were screened and 120 participants were included in the study sample (**Figure 3**). Two hundred and seven participants (41.5%) were ineligible because they had chronic LBP, two participants were excluded because they had previous spinal surgery and three were excluded because physical examination suggested the presence of lumbosacral radiculopathy. Of the 286 eligible participants, 94 (32.9%) failed to respond to contact attempts organising baseline assessment and 72 (25.2%) declined participation after reviewing the study information sheet.

**Figure 3.**
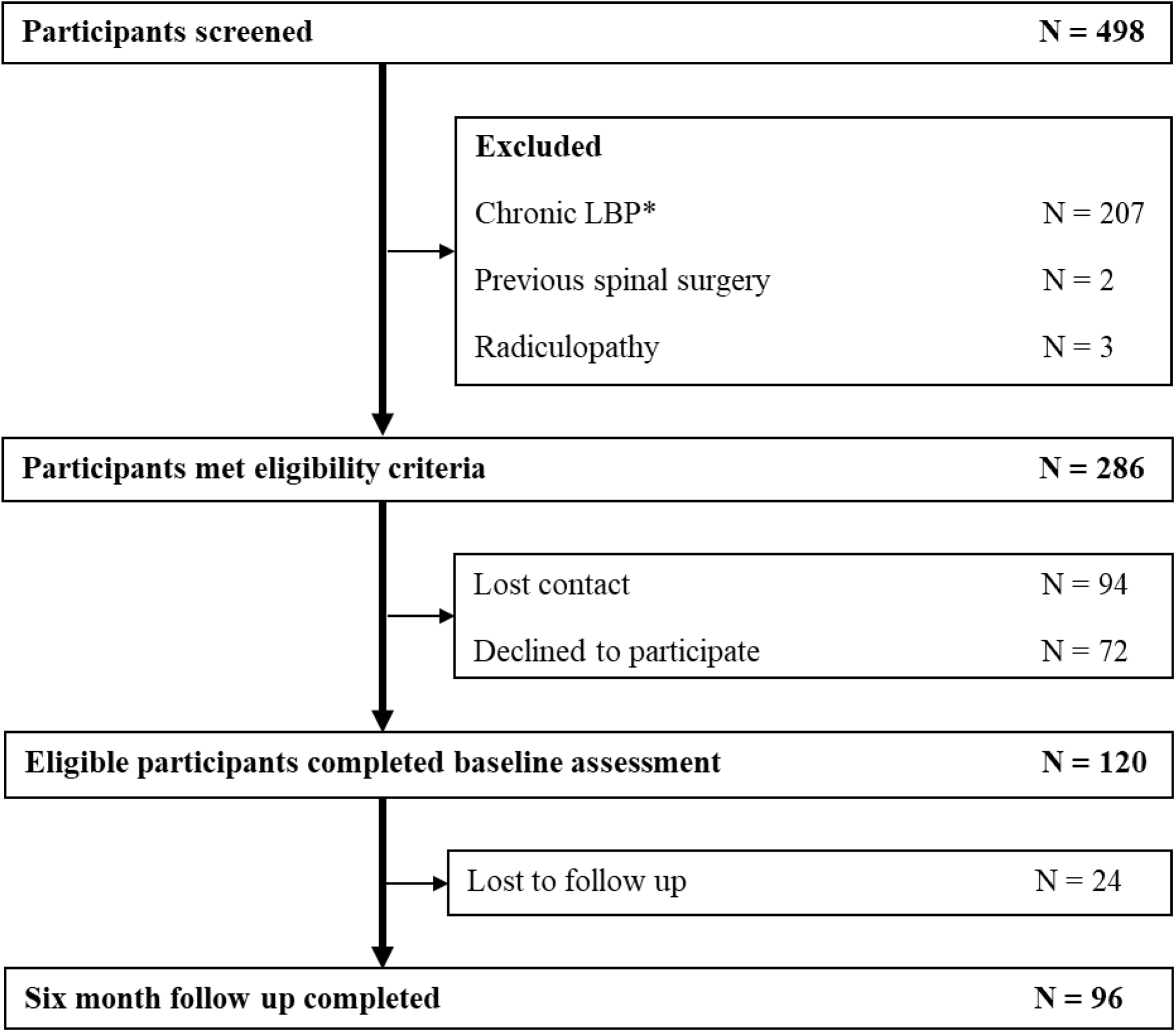
Study flow chart. *defined as LBP lasting for longer than six weeks’ duration and/or LBP episode preceded by a period of less than one-month pain-free.

Follow-up at six months was completed in 96 participants (80%). Missing follow-up cases were due to the participant failing to respond to multiple contact attempts. In the complete case analysis, 67 participants (70%) had a NRS score of ≥ 1 and 35 (36%) had a RMDQ score ≥ 3. Following multiple imputation, 87 participants (73%) had a NRS score of ≥ 1 and 47 (39%) had a RMDQ score ≥ 3 and were considered to have developed chronic pain or chronic disability.

### 3.2. Acute-stage sensorimotor cortex excitability is lower in those who develop chronic pain

Between group differences for somatosensory cortex and corticomotor excitability are presented in **Table 1** (chronic pain) and **Table 2** (chronic disability). The N_80_ SEP area during the acute stage of LBP was smaller in participants who developed chronic pain (*P*_FDR_ = < 0.001) and a similar finding was observed for the N_150_ SEP area (*P*_FDR_ = < 0.001), (**Figure 4)**. There were no between-group differences for N_80_ and N_150_ SEP area during acute LBP when outcome was defined by chronic disability (**Figure 5**). Map volume at the L3 recording site was smaller during the acute stage of LBP in participants who developed chronic pain (*P*_FDR_ = 0.01, **Figure 6**) and in participants who developed chronic disability (*P*_FDR_ = 0.03, **Figure 7**). Map volume at the L5 recording site did not differ between groups.

**Table 1.** Baseline characteristics of participants when outcome is defined by perceived pain at 6-month follow-up. Chronic pain (N = 87) was defined by the presence of pain (NRS ≥ 1) and recovery by the absence of pain (N = 33, NRS=0) at six-month follow up.

| Characteristic | Recovered (N = 33) | Chronic pain (N = 87) | $P_{FDR}$ value |
| --- | --- | --- | --- |
| Gender: Female (%) | 46.9 | 50.0 | 0.92 |
| Previous history of LBP: No (%) | 25.0 | 19.3 | 0.92 |
| BDNF genotype: AA/AG (%) | 31.3 | 43.2 | 0.54 |
| Cultural diversity: No (%) | 50.0 | 52.3 | 0.92 |
| L3 map volume (cm<sup>2</sup>) | 10.4 (3.7) | 8.1 (4.0) | **0.01** |
| L5 map volume (cm<sup>2</sup>) | 8.4 (2.8) | 7.9 (3.4) | 0.66 |
| Log-transformed N<sub>80</sub> SEP area (μV) | -1.6 (1.4) | -3.3 (1.4) | **< 0.001** |
| Log-transformed N<sub>150</sub> SEP area (μV) | -1.6 (1.2) | -3.1 (1.3) | **< 0.001** |
| Age (years) <sup>\$</sup> | 34.3 (11.9) | 41.7 (15.4) | **0.02** |
| Socioeconomic status (SEIFA score) | 1023.5 (51.1) | 1025.3 (61.2) | 0.92 |
| BDNF serum concentration (pg/mL) | 47946.2 (11919.4) | 52517.5 (12987.0) | 0.15 |
| Log-transformed CRP (pg/mL) | 14.2 (1.2) | 14.7 (1.2) | 0.13 |
| TNF (pg/mL) | 6.7 (1.6) | 7.7 (1.9) | **0.02** |
| PCS score <sup>\$</sup> | 6.2 (6.9) | 12.7 (10.4) | **0.001** |
| DASS-21 score <sup>\$</sup> | 10.4 (11.4) | 24.3 (21.2) | **< 0.001** |
| PSEQ score <sup>\$</sup> | 52.8 (8.2) | 43.8 (12.3) | **< 0.001** |
| Local sensitivity (kPa) | 914.2 (355.4) | 868.6 (353.3) | 0.69 |
| Distal sensitivity (kPa) | 639.6 (261.4) | 679.3 (244.0) | 0.66 |

Variable means were compared between non-recovered and recovered low back pain participants using t tests (continuous variable) or $\chi^2$ tests (categorical variables).
<sup>\$</sup> Welch's t test was performed.
Statistically significant values are in bold font
Continuous data described as pooled mean ± pooled SD. Categorical data described as percent.
AA/AG - G allele encodes Val, A allele encodes Met; BDNF – brain derived neurotrophic factor; CRP - C-reactive protein; DASS – depression, anxiety, stress subscale; FDR – false discover rate; LBP – low back pain; PCS – pain catastrophizing scale; PSEQ – pain self-efficacy questionnaire; TNF-α - tumor necrosis factor- $\alpha$ .

**Table 2.** Baseline characteristics of participants when outcome is defined by disability. Chronic disability (N = 47) was defined by RMDQ ≥ 3 and recovery by RMDQ score of ≤ 2 (N = 73) at six-month follow up.

| Characteristic | Recovered (N = 73) | Chronic disability (N = 47) | $P_{FDR}$ value |
| --- | --- | --- | --- |
| Gender: Female (%) | 52.1 | 44.9 | 0.62 |
| Previous history of LBP: No (%) | 23.9 | 16.3 | 0.38 |
| BDNF genotype: AA/AG (%) | 45.1 | 32.7 | 0.38 |
| Cultural diversity: No (%) | 49.3 | 55.1 | 0.57 |
| L3 map volume (cm<sup>2</sup>) | 9.5 (3.8) | 7.5 (4.1) | **0.03** |
| L5 map volume (cm<sup>2</sup>) | 8.3 (3.1) | 7.7 (3.6) | 0.45 |
| Log-transformed N<sub>80</sub> SEP area (μV) | -2.6 (1.5) | -3.2 (1.5) | 0.11 |
| Log-transformed N<sub>150</sub> SEP area (μV) | -2.6 (1.5) | -2.9 (1.5) | 0.32 |
| Age (years) <sup>\$</sup> | 36.8 (13.4) | 43.9 (16.0) | **0.04** |
| Socioeconomic status (SEIFA score) | 1021.3 (58.4) | 1030.1 (58.8) | 0.54 |
| BDNF serum concentration (pg/mL) | 49478.9 (12394.7) | 53935.0 (13100.9) | 0.15 |
| Log-transformed CRP (pg/mL) | 14.4 (1.1) | 14.7 (1.3) | 0.28 |
| TNF (pg/mL) | 7.2 (1.8) | 7.8 (1.9) | 0.15 |
| PCS score <sup>\$</sup> | 8.3 (8.5) | 14.9 (10.7) | **< 0.001** |
| DASS-21 score <sup>\$</sup> | 14.1 (16.1) | 30.1 (21.5) | **< 0.001** |
| PSEQ score <sup>\$</sup> | 49.3 (10.6) | 41.7 (12.5) | **< 0.001** |
| Local sensitivity (kPa) | 868.8 (346.0) | 898.0 (365.7) | 0.70 |
| Distal sensitivity (kPa) | 661.8 (260.3) | 678.8 (232.1) | 0.71 |

Variable means were compared between non-recovered and recovered low back pain participants using t tests (continuous variable) or $\chi^2$ tests (categorical variables).
<sup>\$</sup> Welch's t test was performed.
Statistically significant values are in bold font
Continuous data described as pooled mean ± pooled SD. Categorical data described as percent.
AA/AG - G allele encodes Val, A allele encodes Met; BDNF – brain derived neurotrophic factor; CRP - C-reactive protein; DASS – depression, anxiety, stress subscale; FDR – false discover rate; LBP – low back pain; PCS – pain catastrophizing scale; PSEQ – pain self-efficacy questionnaire; TNF-α - tumor necrosis factor- $\alpha$ .

**Figure 4.**
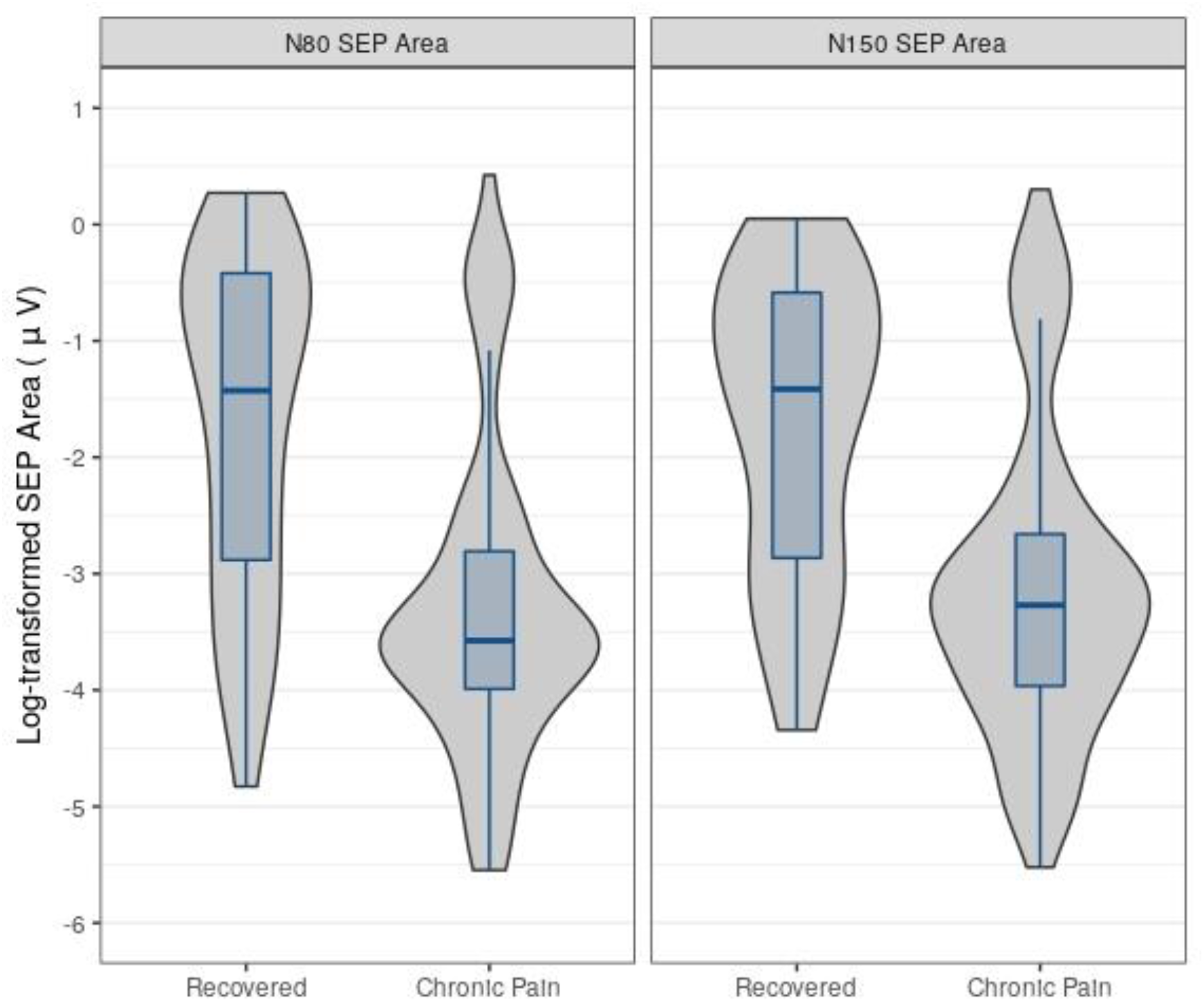
Violin plots displaying the log-transformed distribution of baseline primary (N_80_) and secondary (N_150_) sensory evoked potential area under the curve mean amplitude (µV) values, divided into those who recovered from their episode of acute LBP and those who developed chronic pain (NRS ≥ 1) at six-month follow-up. Boxplots represent median (horizontal line), 25th and 75th percentiles (box), and 10^th^ and 90th percentiles (lines outside the box). Raw values were log-transformed prior to analysis.

**Figure 5.**
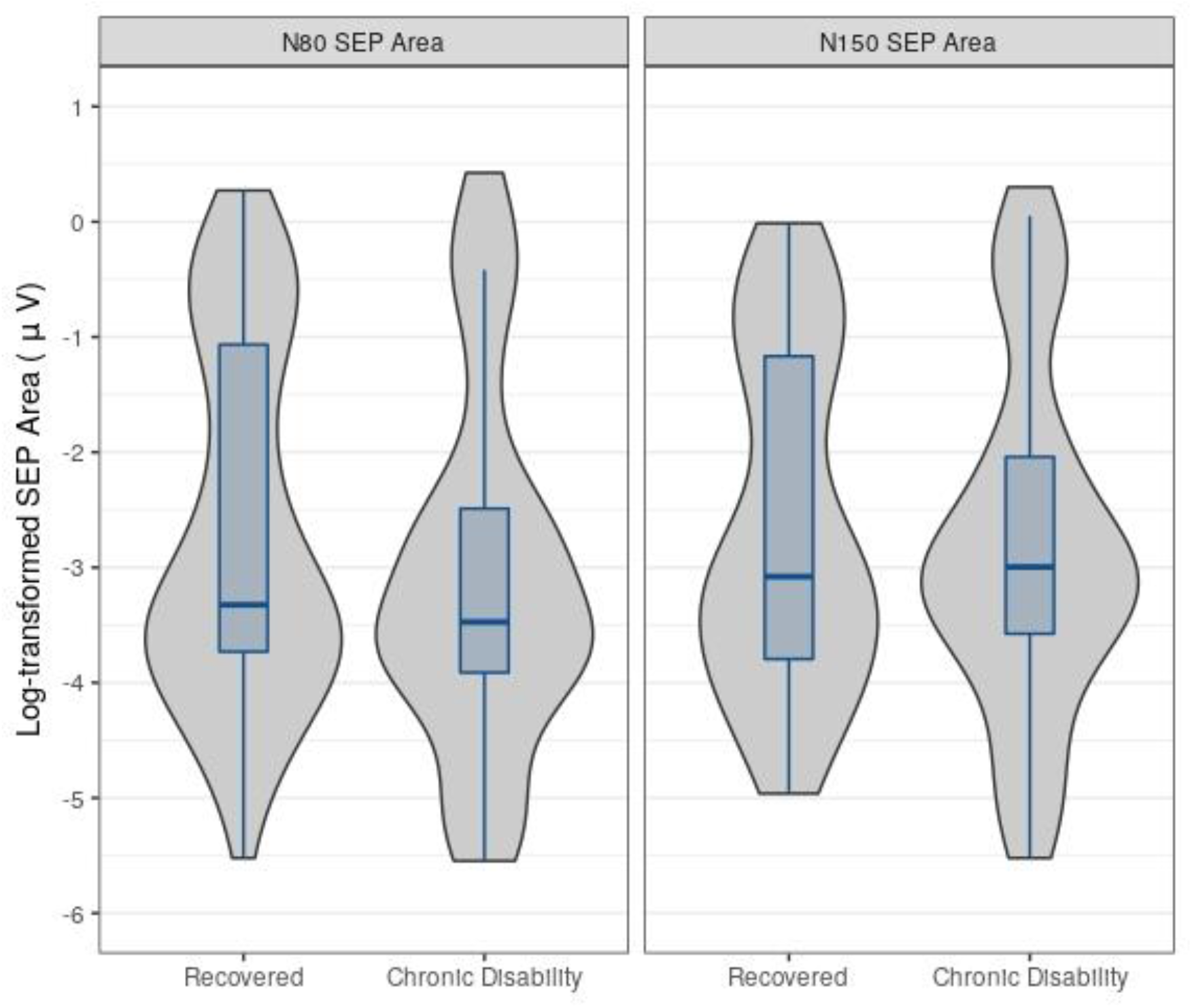
Violin plots displaying the log-transformed distribution of baseline primary (N_80_) and secondary (N_150_) sensory evoked potential area under the curve mean amplitude (µV) values, divided into those who recovered from their episode of acute LBP and those who developed **chronic disability** (RMDQ ≥ 3) at six-month follow-up. Boxplots represent median (horizontal line), 25th and 75th percentiles (box), and 10^th^ and 90th percentiles (lines outside the box). Raw values were log-transformed prior to analysis.

**Figure 6.**
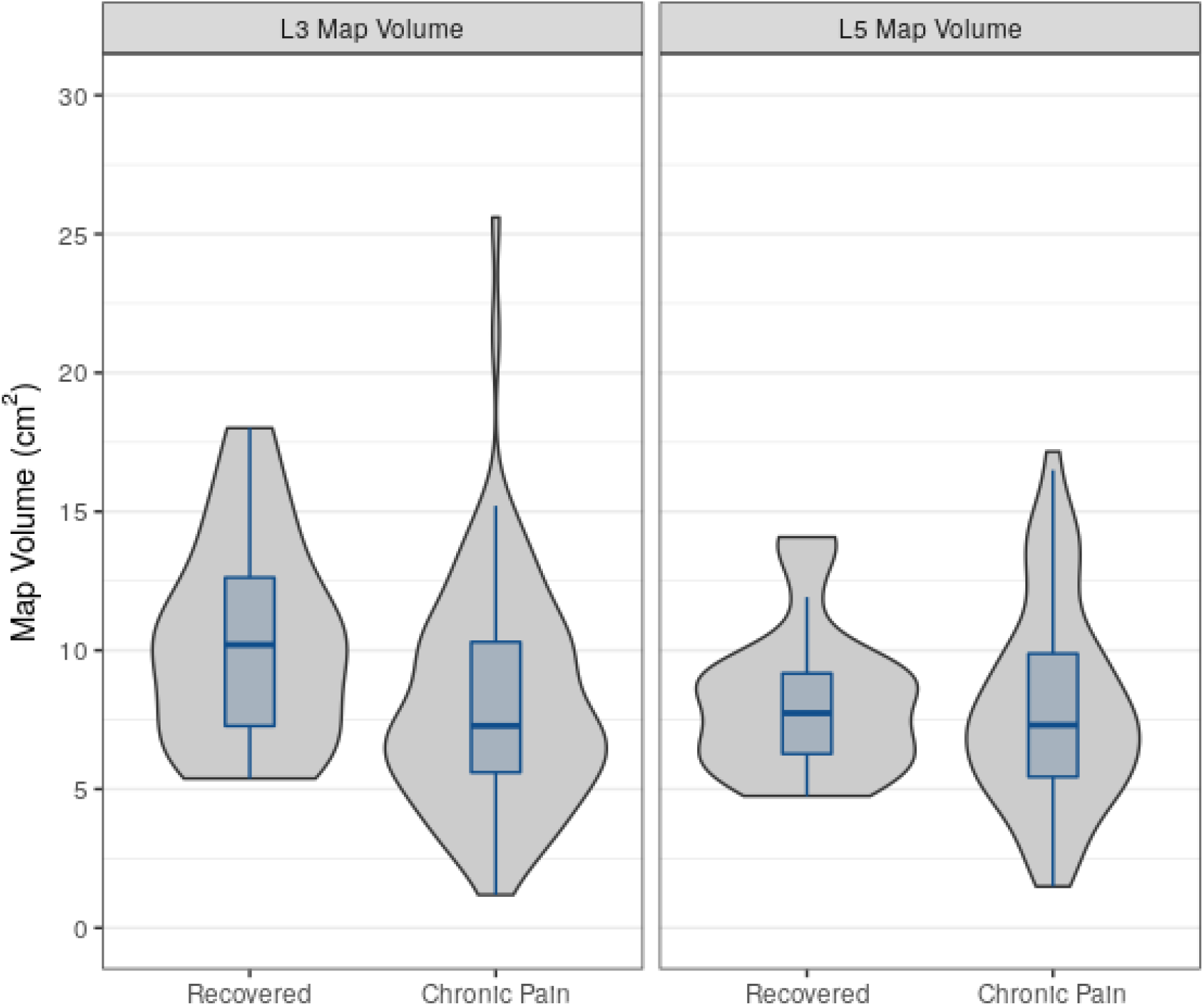
Violin plots displaying the distribution of baseline map volume (cm^2^) from the L3 and L5 electromyographic recording sites, divided into those who recovered from their episode of acute LBP and those who developed **chronic pain** (NRS ≥1) at six-month follow-up. Boxplots represent median (horizontal line), 25th and 75th percentiles (box), and 10^th^ and 90th percentiles (lines outside the box).

**Figure 7.**
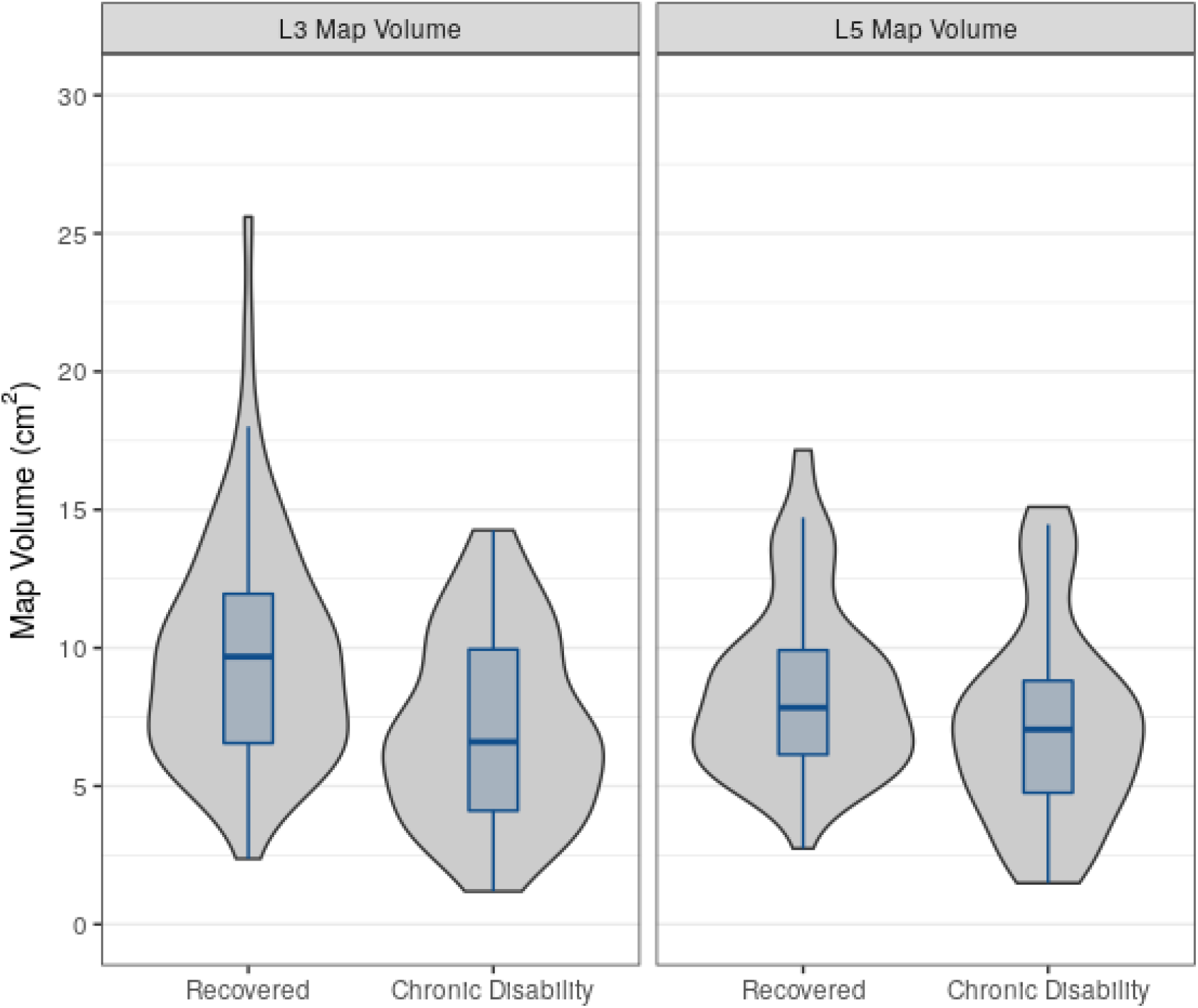
Violin plots displaying the distribution of baseline map volume (cm^2^) from the L3 and L5 electromyographic recording sites, divided into those who recovered from their episode of acute LBP and those who developed **chronic disability** (RMDQ ≥ 3) at six-month follow-up. Boxplots represent median (horizontal line), 25th and 75th percentiles (box), and 10^th^ and 90th percentiles (lines outside the box).

### 3.3. Older age, higher serum concentrations of TNF-α and pain-related psychological status in the acute stage of LBP are associated with chronic pain and chronic disability

**Table 1** and **Table 2** also present baseline characteristics for confounding variables. Older participants were more likely to develop chronic pain (*P*_FDR_ = 0.02) and chronic disability (*P*_FDR_ = 0.04). Amongst the blood biomarkers analysed, participants who developed chronic pain had higher serum concentrations of TNF-α (*P*_FDR_ = 0.02) in the acute stage of LBP, but this was not observed when the outcome was chronic disability. Participants who developed chronic pain or chronic disability demonstrated lower levels of pain self-efficacy, higher DASS-21 scores and higher PCS scores (*P*_FDR_ = < 0.01). No between-group differences were observed for any other variable.

### 3.4. Lower acute-stage somatosensory cortex excitability increases the likelihood of developing chronic pain but not chronic disability

The effects of acute-stage somatosensory cortex excitability on six-month LBP outcome are described in **Table 3** and **Table 4**. Acute-stage N_80_ (log-transformed; B = −0.56, 95% CI: −0.82 to −0.30, *P*_FDR_ = < 0.001) and N_150_ (log-transformed; B = −0.52, 95% CI: −0.81 to −0.23, *P*_FDR_ = 0.001) SEP area were associated with six-month pain intensity (continuous variable). After adjustment for confounding, N_80_ (B = −0.31, 95% CI: −0.60 to −0.03, *P*_FDR_ = 0.06) and N_150_ (B = −0.36, 95% CI = −0.66 to −0.07, *P*_FDR_ = 0.06) SEP area in the acute stage of LBP demonstrated an inverse relationship with six-month pain intensity. However, this effect did not reach statistical significance following FDR correction.

**Table 3.** Adjusted and unadjusted linear regression models to test the effects of baseline sensorimotor cortex activity on pain intensity and RMDQ score at six-month follow-up.

| Outcome | Model | Exposure | B (95% CI) | $P_{FDR}$ |
| --- | --- | --- | --- | --- |
| Pain Intensity | Unadjusted | Log-transformed N<sub>80</sub> SEP area | -0.56 (-0.82, -0.30) | < **0.001** |
|  |  | Log-transformed N<sub>150</sub> SEP area | -0.52 (-0.81, -0.23) | **0.001** |
|  |  | L3 map volume | -0.15 (-0.28, -0.02) | **0.03** |
|  |  | L5 map volume | -0.06 (-0.20, 0.08) | 0.38 |
|  | Adjusted<sup>1</sup> | Log-transformed N<sub>80</sub> SEP area | -0.31 (-0.60, -0.03) | 0.06 |
|  |  | Log-transformed N<sub>150</sub> SEP area | -0.36 (-0.66, -0.07) | 0.06 |
|  |  | L3 map volume | -0.11 (-0.23, 0.02) | 0.14 |
|  |  | L5 map volume | -0.02 (-0.15, 0.11) | 0.80 |
| RMDQ score | Unadjusted | Log-transformed N<sub>80</sub> SEP area | -0.35 (-0.95, 0.25) | 0.56 |
|  |  | Log-transformed N<sub>150</sub> SEP area | -0.26 (-0.91, 0.38) | 0.56 |
|  |  | L3 map volume | -0.13 (-0.41, 0.15) | 0.56 |
|  |  | L5 map volume | 0.01 (-0.33, 0.35) | 0.96 |
|  | Adjusted<sup>1</sup> | Log-transformed N<sub>80</sub> SEP area | -0.24 (-0.84, 0.36) | 0.58 |
|  |  | Log-transformed N<sub>150</sub> SEP area | -0.34 (-0.93, 0.25) | 0.58 |
|  |  | L3 map volume | -0.12 (-0.37, 0.13) | 0.58 |
|  |  | L5 map volume | 0.01 (-0.29, 0.31) | 0.92 |

<sup>1</sup> Adjusted for predisposing factors, blood biomarkers, psychological variables and sensitisation.
Statistically significant values are in bold font
B – unstandardized beta coefficient, CI – confidence interval, FDR – false discovery rate, SEP – sensory evoked potential

**Table 4.**
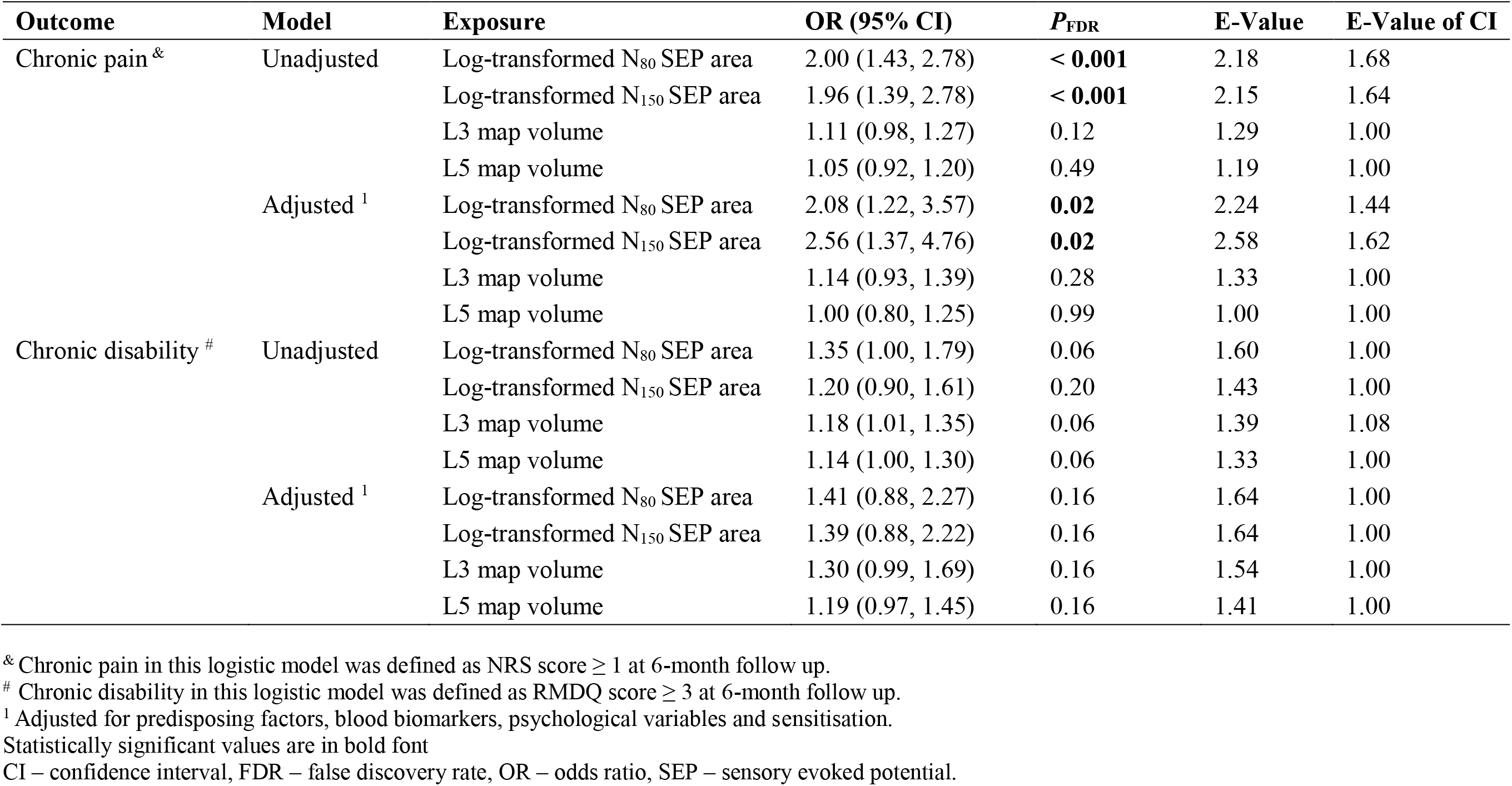
Adjusted and unadjusted logistic regression models to test the effects of baseline sensorimotor activity on chronic pain (NRS ≥ 1) and chronic disability (RMDQ ≥ 3) at six-month follow-up.

| Outcome | Model | Exposure | OR (95% CI) | $P_{\text{FDR}}$ | E-Value | E-Value of CI |
| --- | --- | --- | --- | --- | --- | --- |
| Chronic pain <sup>&</sup> | Unadjusted | Log-transformed N<sub>80</sub> SEP area | 2.00 (1.43, 2.78) | **< 0.001** | 2.18 | 1.68 |
|  |  | Log-transformed N<sub>150</sub> SEP area | 1.96 (1.39, 2.78) | **< 0.001** | 2.15 | 1.64 |
|  |  | L3 map volume | 1.11 (0.98, 1.27) | 0.12 | 1.29 | 1.00 |
|  |  | L5 map volume | 1.05 (0.92, 1.20) | 0.49 | 1.19 | 1.00 |
|  | Adjusted <sup>1</sup> | Log-transformed N<sub>80</sub> SEP area | 2.08 (1.22, 3.57) | **0.02** | 2.24 | 1.44 |
|  |  | Log-transformed N<sub>150</sub> SEP area | 2.56 (1.37, 4.76) | **0.02** | 2.58 | 1.62 |
|  |  | L3 map volume | 1.14 (0.93, 1.39) | 0.28 | 1.33 | 1.00 |
|  |  | L5 map volume | 1.00 (0.80, 1.25) | 0.99 | 1.00 | 1.00 |
| Chronic disability <sup>#</sup> | Unadjusted | Log-transformed N<sub>80</sub> SEP area | 1.35 (1.00, 1.79) | 0.06 | 1.60 | 1.00 |
|  |  | Log-transformed N<sub>150</sub> SEP area | 1.20 (0.90, 1.61) | 0.20 | 1.43 | 1.00 |
|  |  | L3 map volume | 1.18 (1.01, 1.35) | 0.06 | 1.39 | 1.08 |
|  |  | L5 map volume | 1.14 (1.00, 1.30) | 0.06 | 1.33 | 1.00 |
|  | Adjusted <sup>1</sup> | Log-transformed N<sub>80</sub> SEP area | 1.41 (0.88, 2.27) | 0.16 | 1.64 | 1.00 |
|  |  | Log-transformed N<sub>150</sub> SEP area | 1.39 (0.88, 2.22) | 0.16 | 1.64 | 1.00 |
|  |  | L3 map volume | 1.30 (0.99, 1.69) | 0.16 | 1.54 | 1.00 |
|  |  | L5 map volume | 1.19 (0.97, 1.45) | 0.16 | 1.41 | 1.00 |

<sup>&</sup> Chronic pain in this logistic model was defined as NRS score $\geq 1$ at 6-month follow up.
<sup>#</sup> Chronic disability in this logistic model was defined as RMDQ score $\geq 3$ at 6-month follow up.
<sup>1</sup> Adjusted for predisposing factors, blood biomarkers, psychological variables and sensitisation.
Statistically significant values are in bold font
CI – confidence interval, FDR – false discovery rate, OR – odds ratio, SEP – sensory evoked potential.

Logistic regression models showed smaller N_80_ SEP area in the acute stage of LBP increased the likelihood of developing chronic pain (dichotomised variable) after adjustment for confounding variables (OR = 2.08, 95% CI = 1.22 to 3.57, *P*_FDR_ = 0.02). Similarly, a smaller N_150_ SEP area at baseline increased the likelihood of developing chronic pain after adjustment for confounding variables (OR = 2.56, 95% CI = 1.37 to 4.76, *P*_FDR_ = 0.02). No effect of somatosensory cortex excitability on chronic disability was observed after adjustment for confounding.

### 3.5. Lower acute-stage corticomotor excitability does not increase the likelihood of developing chronic pain or disability

The effect of acute-stage corticomotor excitability on six-month LBP outcome is described in **Table 3** and **Table 4**. An association between map volume at the L3 recording site and six-month pain intensity was observed (B = −0.15, 95% CI: −0.28 to −0.02, *P*_FDR_ = 0.03), however, this effect did not remain after adjustment for confounding. Thus, L3 map volume demonstrated no causal effect on the development of chronic pain. Map volume at the L5 recording site showed no association with six-month pain intensity (continuous variable) or chronic pain (dichotomised variable). No statistically significant association was observed between L3 map volume (OR = 1.18, 95% CI: 1.01 to 1.35, *P*_FDR_ = 0.06) or L5 map volume (OR = 1.14, 95% CI: 1.00 to 1.30, *P*_FDR_= 0.06) and chronic disability (dichotomised variable).

### 3.6. Sensitivity analysis

**Table 4** presents E-Values used to explore the effect of unmeasured confounding. An E-Value of 2.24 (E-Value of CI = 1.44) and 2.58 (E-Value of CI = 1.62) were calculated for the log-transformed N_80_ and N_150_ SEP area respectively, providing plausible evidence for a true causal effect. For example, for the N_80_ SEP area, an unmeasured confounder would have to be associated with both the exposure and outcome by a risk ratio of 2.24-fold, through pathways independent of multiple measured confounders (i.e. predisposing factors, blood biomarkers, psychological variables and sensitisation) to explain away the observed effect. Smaller E-Values were observed for corticomotor excitability variables suggesting greater sensitivity to unmeasured confounding.

As shown in Appendix 1, the results of the complete cases analysis mirror that of the imputed data. Linear regression models demonstrated the same statistically significant effects following FDR correction. Logistic regression models demonstrated ORs and corresponding confidence intervals similar to the imputed data. The loss of effect for adjusted N_80_ and N_150_ SEP area measures on chronic pain development following FDR correction suggests the reduction in sample size decreased statistical power to detect an effect. In combination, these findings suggest missing data did not impact the results.

## 4. Discussion

This study is the first prospective, longitudinal, cohort study to investigate neurophysiological mechanisms underpinning the transition from acute to chronic LBP using rigorous methods from the causal inference field (30). Our findings demonstrate that a smaller N_80_ and N_150_ SEP area, reflecting lower somatosensory cortex excitability, during an acute episode of LBP increases the likelihood that an individual will develop chronic pain. This effect remains after adjustment for predisposing factors, blood biomarkers, psychological variables and pressure pain sensitivity, suggesting the observed effect is robust to confounding bias. Thus, lower somatosensory cortex excitability during the acute stage of LBP may represent a physiologically relevant causal mechanism underpinning the development of chronic LBP.

Previous research has identified brain regions responsible for processing acute pain including the thalamus, primary and secondary somatosensory cortices, insular and anterior cingulate cortex (S1, S2, IC, ACC) (90). Brain responses to painful stimuli in healthy, pain-free people result in transmission of afferent nociceptive information via spinothalamic pathways to the thalamus, S1, S2, IC and ACC. When painful stimuli are processed by people suffering chronic pain these brain areas decrease in activation incidence (90). In the current study, lower S1 and S2 excitability in response to non-noxious stimuli applied to the painful lumbar region during acute LBP contributed to the development of chronic LBP.

During chronic pain, the decreased activation incidence of thalamus, S1, S2, IC and ACC is contrasted by increased activation incidence of the pre-frontal cortex (PFC) compared with pain processing in healthy, pain-free individuals (90-92). It is widely agreed that chronic pain states have stronger affective, motivational, and cognitive components and this underpins the preferential activation of the PFC (90, 93). Whilst activity within the PFC was not directly measured in our study, psychological functions associated with these regions were. People who developed chronic LBP had higher levels of depression, anxiety and stress, higher levels of pain catastrophizing and lower levels of pain self-efficacy during their acute LBP episode than those who recovered. The tendency to have a stronger affective response to an acute episode of LBP may be a result of decreased activation of brain regions such as S1 and S2 and a shift in activation towards PFC regions (17). Previous research provides support for this theory, suggesting white-matter network connectivity between dorsal medial PFC, amygdala and nucleus accumbens during sub-acute LBP accounts for 60% of the variance in chronic LBP development at three year follow up (94).

Through the causal analyses of longitudinal, observational data undertaken here, we provide empirical evidence for these existing theories, and generate new hypotheses for potential treatment targets (95). Specifically, our data suggest low excitability within somatosensory regions may be a key causal factor underpinning the development of chronicity. These unique findings suggest interventions designed to elevate somatosensory cortex excitability in the acute stage of pain may interfere with the development of chronic pain, providing a new area of research focus. For example, excitatory repetitive transcranial magnetic stimulation (96), and sensorimotor retraining (97) are promising, non-invasive treatments, that could be optimised to target low somatosensory cortex excitability in the acute stage of LBP. Exploring interventions that can target the neurophysiological mechanisms of chronic LBP is particularly important given interventions that influence the psychosocial aspects of pain processing such as intensive education or cognitive behavioural therapy, do not seem sufficient to prevent or treat chronic LBP when applied in isolation (4, 98).

Baseline corticomotor excitability demonstrated an association with six-month pain intensity. Participants with higher six-month pain intensity had a smaller M1 map volume at the L3 recording site, but not the L5 recording site, in the acute stage of LBP. Smaller map volume of the paraspinal extensor muscles, indicating reduced corticomotor excitability, has been observed previously in people with acute (16), and chronic LBP (23) when compared to healthy controls. Previous studies have suggested reduced corticomotor excitability may represent an attempt to limit provocative movements thus minimising the threat (actual or potential) of further pain and injury (99, 100). It is hypothesised this could reduce short-term pain intensity but contribute to long-term consequences, including increased load on spinal structures, long-term reductions in movement, and decreased movement variability (99). While our data provide some support for this theory, adjustment for the sufficient set of confounding variables removed the observed association between acute-stage corticomotor excitability and six-month pain intensity, suggesting other factors present during acute-stage LBP are more likely to cause chronic LBP. Appropriate consideration of confounding is essential in observational studies attempting to draw causal inferences, thus this finding should be considered a non-causal association (29, 30). The finding that somatosensory cortex excitability had a causal effect on chronic LBP development, whilst corticomotor activity did not, is not entirely unexpected. Previous research has suggested S1 and M1 act independently during pain processing (101). During acute, experimental muscle pain, reduced S1 excitability occurs prior to reduced motor output, via processes that are non-linear and involve longer information processing times (101). Our data provide further support for a complex and independent relationship between somatosensory and corticomotor excitability during pain.

This study is not without limitations. First, although missing data are inevitable in longitudinal trials, the presence of incomplete cases represents a threat to the validity of the results. To control for this, missing values were imputed (102). The multiple imputation procedure used in this study is thought to produce the least biased regression coefficient estimates and is recommended for use in practice (103). A sensitivity analysis (Appendix 1) demonstrated no difference between regression models with and without imputed data, suggesting the impact of missing data on the study results was negligible. Second, although the use of DAGs is the only approach to confounder adjustment that make causal assumptions explicit and transparent (28, 30), DAG development is reliant on expert content knowledge and may not include unknown/unmeasured confounders (28). The DAG used to develop the causal model for this study is publicly available and can be used as a foundation for future research (34). Further, in line with recommendations from the STROBE statement, we performed and reported a sensitivity analysis to explore the effect of unmeasured confounding. As E-values were large in the current study for confounder-adjusted estimates of S1 and S2 effect sizes, unmeasured confounding acting through pathways that were not controlled, is unlikely to explain the causal effect of acute-stage somatosensory cortex excitability on chronic pain development (89, 104, 105). Finally, S1 and S2 activity may have been lower in a pre-pain state for the participants who developed chronic pain, representing a predisposition to chronic pain. This cannot be elucidated from the current study.

## 5. CONCLUSION

This study provides novel evidence that low somatosensory cortex excitability in the acute stage of LBP causes chronic pain. Whilst this study identifies an inverse association between acute-stage corticomotor activity and six-month pain intensity no causal effect was observed after confounder adjustment. Future research should confirm the causal interpretation of this study with interventions designed to elevate somatosensory cortex excitability in the acute stage of LBP using randomized controlled study designs. Future research should also seek to identify other physiological and biological causal mechanisms underpinning chronic pain development and expand upon the causal model of chronic LBP developed in this study.

## Data Availability

The data that support the findings of this study are available on request from the corresponding author, [SMS]. The data are not publicly available to ensure the privacy of research participants.

## Acknowledgements

We thank the patients who participated in the study. We thank Dr Valerie Wasinger, Biosciences Precinct Laboratory, Level 2, Biosciences South (E26), UNSW Sydney, NSW 2052 for her expertise and assistance with serum analyses.

