## Appendix 1 for "Low somatosensory cortex excitability in the acute stage of low back pain causes chronic pain"

**Complete case analysis - Table 1.** Baseline characteristics of participants with complete data when outcome is defined by perceived pain at 6-month follow-up. Chronic pain (N = 67) was defined by the presence of pain (NRS ≥ 1) and recovery by the absence of pain (N = 29, NRS=0) at six-month follow up.

| **Characteristic** | **Recovered (N = 29)** | **Chronic pain (N = 67)** | $P_{FDR}$ **value** |
|---|---|---|---|
| Gender: Female (%) | 48.2 | 55.2 | 0.88 |
| Previous history of LBP: No (%) | 28.6 | 21.2 | 0.86 |
| BDNF genotype: AA/AG (%) | 31.0 | 38.8 | 0.86 |
| Cultural diversity: No (%) | 53.6 | 59.1 | 0.95 |
| L3 map volume ($cm^2$) | 9.6 (3.8) | 7.9 (4.0) | 0.22 |
| L5 map volume ($cm^2$) | 7.8 (2.6) | 7.9 (3.9) | 0.96 |
| Log-transformed $N_{80}$ SEP area (µV) | -1.6 (1.3) | -3.2 (1.4) | **< 0.001** |
| Log-transformed $N_{150}$ SEP area (µV) | -1.6 (1.2) | -3.0 (1.4) | **< 0.001** |
| Age (years) $^{\$}$ | 33.9 (11.9) | 42.1 (16.4) | **0.02** |
| Socioeconomic status (SEIFA score) | 1023.7 (51.3) | 1021.7 (64.5) | 0.96 |
| BDNF serum concentration (pg/mL) | 48189.3 (11343.6) | 54030.1 (14462.4) | 0.22 |
| Log-transformed CRP (pg/mL) | 14.2 (1.3) | 14.8 (1.4) | 0.22 |
| TNF (pg/mL) | 6.88 (1.8) | 7.8 (2.0) | 0.16 |
| PCS score $^{\$}$ | 6.0 (7.1) | 12.5 (10.9) | **< 0.01** |
| DASS-21 score $^{\$}$ | 8.4 (7.5) | 23.8 (21.0) | **< 0.001** |
| PSEQ score $^{\$}$ | 52.5 (8.5) | 44.8 (12.7) | **< 0.01** |
| Local sensitivity (kPa) | 943.0 (367.1) | 835.0 (361.3) | 0.31 |
| Distal sensitivity (kPa) | 641.3 (269.0) | 638.5 (216.6) | 0.96 |

Variable means were compared between non-recovered and recovered low back pain participants using t tests (continuous variable) or $X^2$ tests (categorical variables).
$^{\$}$ Welch's t test was performed.
Statistically significant values are in bold font
Continuous data described as pooled mean ± pooled SD. Categorical data described as percent.
AA/AG - G allele encodes Val, A allele encodes Met; BDNF – brain derived neurotrophic factor; CRP - C-reactive protein; DASS – depression, anxiety, stress subscale; FDR – false discover rate; LBP – low back pain; PCS – pain catastrophizing scale; PSEQ – pain self-efficacy questionnaire; TNF-α - tumor necrosis factor.

**Complete case analysis - Table 2.** Baseline characteristics of participants with complete data when outcome is defined by disability. Chronic disability (N = 35) was defined by RMDQ ≥ 3 and recovery by RMDQ score of ≤ 2 (N = 61) at six-month follow up.

| **Characteristic** | **Recovered (N = 61)** | **Chronic disability (N = 35)** | $P_{FDR}$ **value** |
|---|---|---|---|
| Gender: Female (%) | 55.7 | 49.0 | 0.71 |
| Previous history of LBP: No (%) | 26.7 | 17.6 | 0.65 |
| BDNF genotype: AA/AG (%) | 41.0 | 28.6 | 0.52 |
| Cultural diversity: No (%) | 55.0 | 61.8 | 0.71 |
| L3 map volume ($cm^2$) | 9.3 (4.0) | 6.6 (3.3) | **0.03** |
| L5 map volume ($cm^2$) | 8.3 (3.3) | 6.9 (3.9) | 0.28 |
| Log-transformed $N_{80}$ SEP area (μV) | -2.4 (1.5) | -3.2 (1.5) | 0.10 |
| Log-transformed $N_{150}$ SEP area (μV) | -2.4 (1.5) | -2.85 (1.47) | 0.34 |
| Age (years) [$] | 36.6 (13.9) | 44.9 (17.0) | 0.06 |
| Socioeconomic status (SEIFA score) | 1019.2 (59.6) | 1027.8 (62.5) | 0.65 |
| BDNF serum concentration (pg/mL) | 50573.7 (13232.0) | 55156.8 (14449.8) | 0.34 |
| Log-transformed CRP (pg/mL) | 14.4 (1.3) | 14.9 (1.4) | 0.29 |
| TNF (pg/mL) | 7.40 (2.0) | 7.8 (1.9) | 0.65 |
| PCS score [$] | 8.0 (8.6) | 15.0 (11.6) | **0.01** |
| DASS-21 score [$] | 14.0 (16.0) | 28.2 (21.6) | **0.01** |
| PSEQ score [$] | 50.6 (9.6) | 40.9 (13.6) | **< 0.01** |
| Local sensitivity (kPa) | 854.9 (352.5) | 887.8 (388.2) | 0.71 |
| Distal sensitivity (kPa) | 641.4 (254.2) | 635.9 (193.4) | 0.91 |

Variable means were compared between non-recovered and recovered low back pain participants using t tests (continuous variable) or $X^2$ tests (categorical variables).
[$] Welch's t test was performed.
Statistically significant values are in bold font
Continuous data described as mean ± SD. Categorical data described as percent.
AA/AG - G allele encodes Val, A allele encodes Met; BDNF – brain derived neurotrophic factor; CRP - C-reactive protein; DASS – depression, anxiety, stress subscale; FDR – false discover rate; LBP – low back pain; PCS – pain catastrophizing scale; PSEQ – pain self-efficacy questionnaire; TNF-α - tumor necrosis factor.

**Complete case analysis - Figure 1.** Violin plots displaying the log-transformed distribution of baseline primary ($N_{80}$) and secondary ($N_{150}$) sensory evoked potential area under the curve mean amplitude (µV) values, divided into those who recovered from their episode of acute LBP and those who developed **chronic pain** (NRS ≥ 1) at 6-months follow up. Boxplots represent median (horizontal line), 25th and 75th percentiles (box), and $10^{th}$ and 90th percentiles (lines outside the box). Raw values were log-transformed prior to analysis.

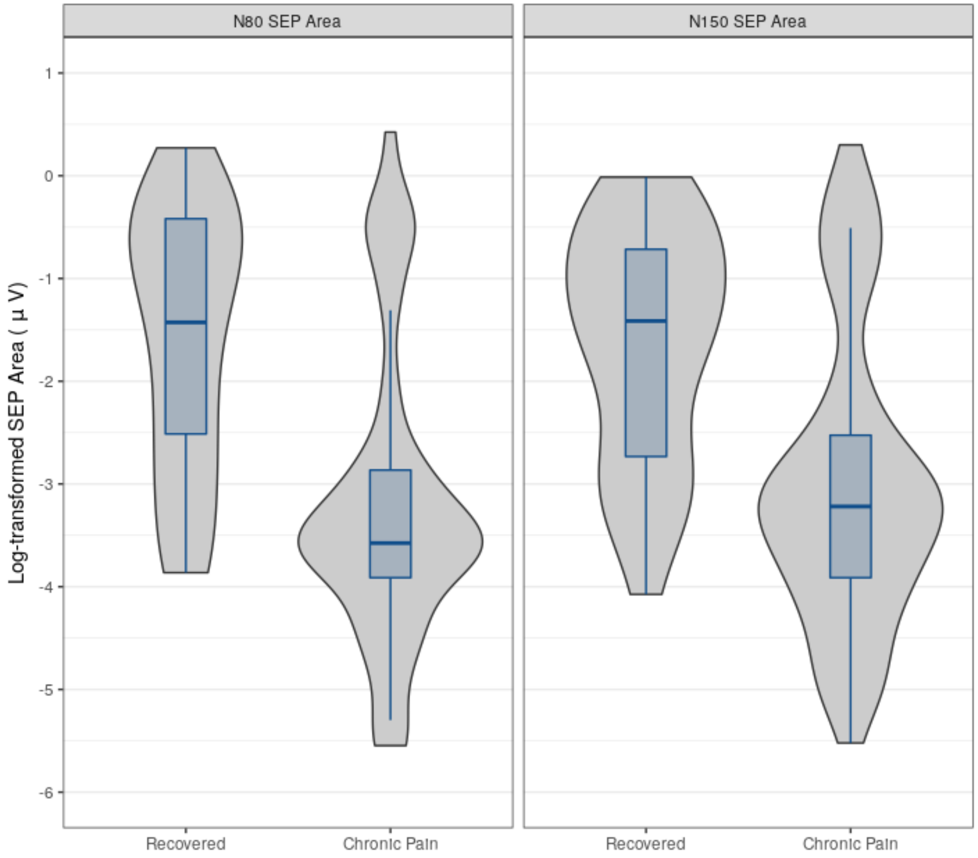

**Complete case analysis - Figure 2.** Violin plots displaying the log-transformed distribution of baseline primary ($N_{80}$) and secondary ($N_{150}$) sensory evoked potential area under the curve mean amplitude (µV) values, divided into those who recovered from their episode of acute LBP and those who developed **chronic disability** (RMDQ ≥ 3) at 6-months follow up. Boxplots represent median (horizontal line), 25th and 75th percentiles (box), and 10$^{th}$ and 90th percentiles (lines outside the box). Raw values were log-transformed prior to analysis.

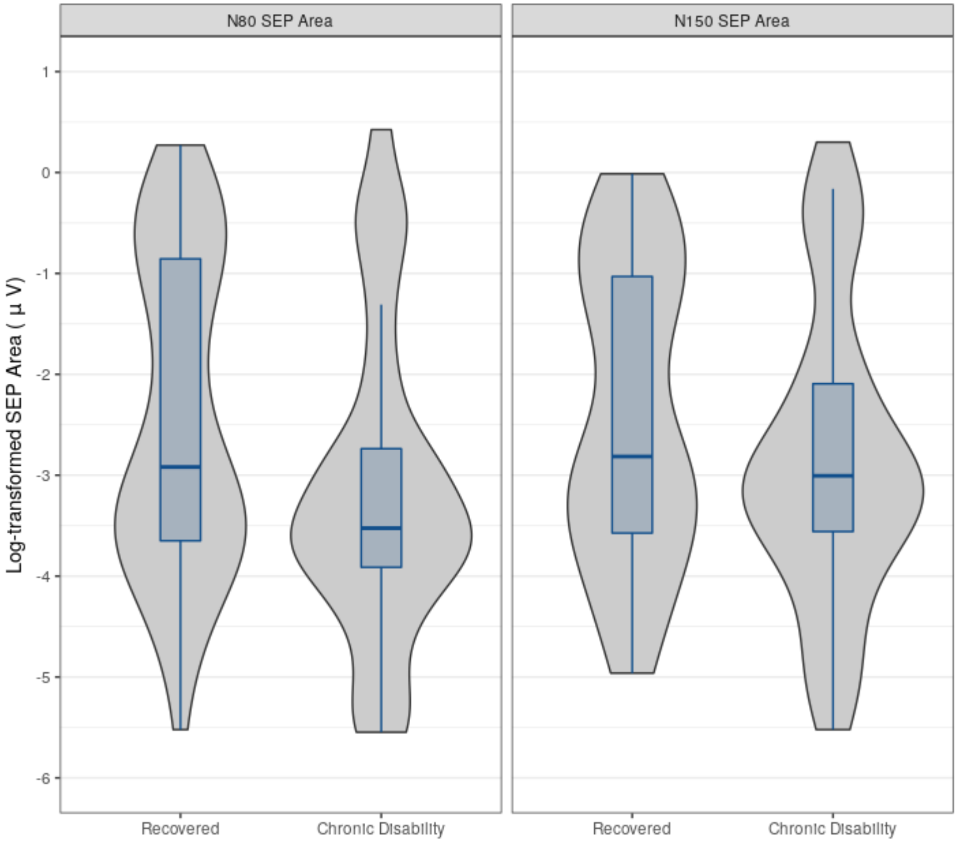

**Complete case analysis - Figure 3.** Violin plots displaying the distribution of baseline map volume ($cm^2$) from the L3 and L5 electromyographic recording sites, divided into those who recovered from their episode of acute LBP and those who developed **chronic pain** (NRS ≥ 1) at six-month follow-up. Boxplots represent median (horizontal line), 25th and 75th percentiles (box), and $10^{th}$ and 90th percentiles (lines outside the box).

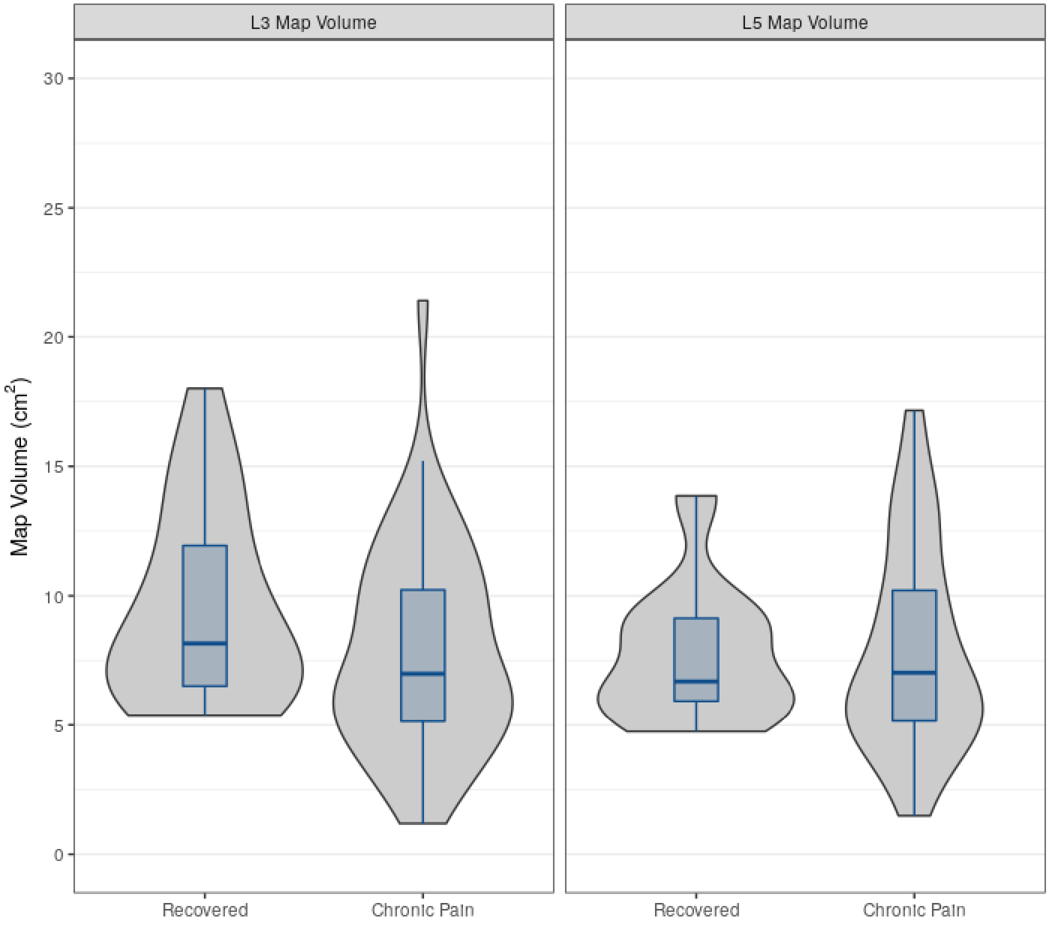

**Complete case analysis - Figure 4.** Violin plots displaying the distribution of baseline map volume (cm$^2$) from the L3 and L5 electromyographic recording sites, divided into those who recovered from their episode of acute LBP and those who developed **chronic disability** (RMDQ $\geq$ 3) at six-month follow-up. Boxplots represent median (horizontal line), 25th and 75th percentiles (box), and 10$^{th}$ and 90th percentiles (lines outside the box).

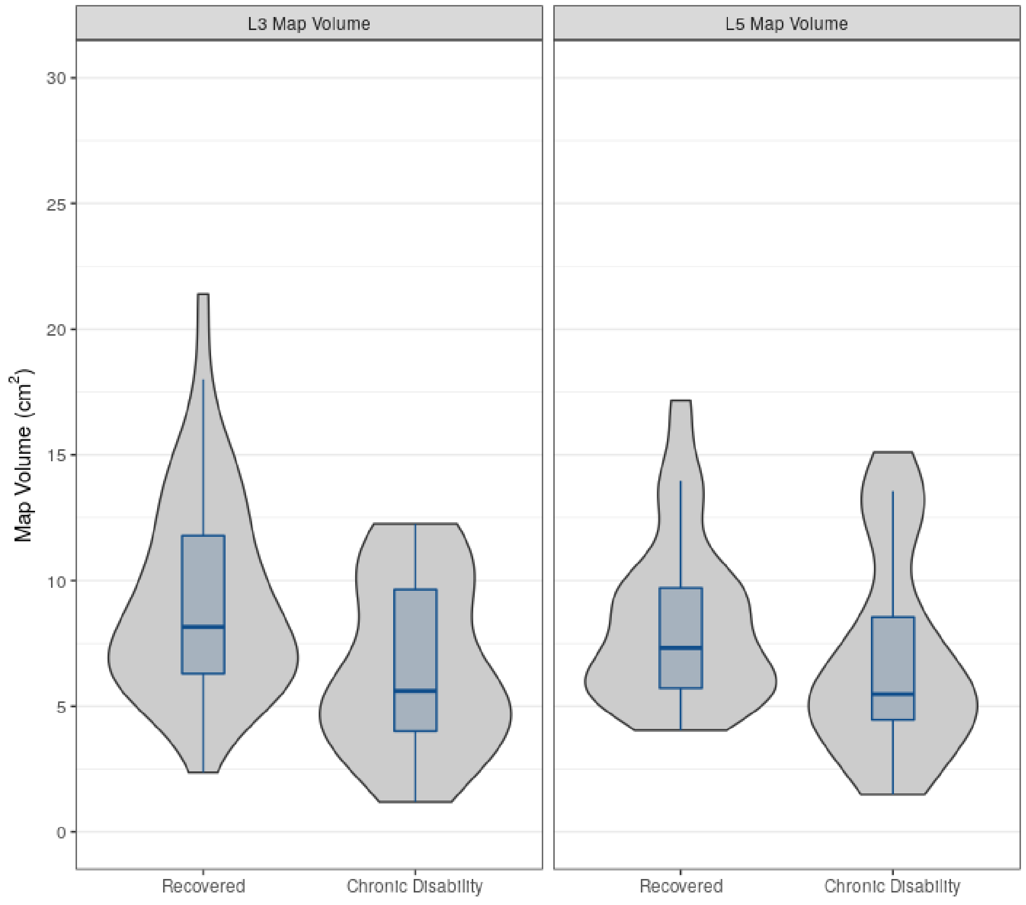

**Complete case analysis - Table 3.** Adjusted and unadjusted linear regression models, including only complete cases, to test the effects of baseline sensorimotor cortex activity on pain intensity and RMDQ score at six-month follow-up.

| Outcome | Model | Exposure | B (95% CI) | $P_{FDR}$ |
|---|---|---|---|---|
| Pain Intensity | Unadjusted | Log-transformed $N_{80}$ SEP area | -0.58 (-0.85, -0.31) | **< 0.001** |
| | | Log-transformed $N_{150}$ SEP area | -0.54 (-0.83, -0.25) | **< 0.01** |
| | | L3 map volume | -0.18 (-0.30, -0.06) | **0.01** |
| | | L5 map volume | -0.06 (-0.20, 0.08) | 0.41 |
| | Adjusted[1] | Log-transformed $N_{80}$ SEP area | -0.14 (-0.55, 0.26) | 0.63 |
| | | Log-transformed $N_{150}$ SEP area | -0.23 (-0.63, 0.17) | 0.49 |
| | | L3 map volume | -0.16 (-0.35, 0.02) | 0.32 |
| | | L5 map volume | -0.03 (-0.22, 0.17) | 0.77 |
| RMDQ score | Unadjusted | Log-transformed $N_{80}$ SEP area | -0.42 (-1.02 ,0.17) | 0.36 |
| | | Log-transformed $N_{150}$ SEP area | -0.33 (-0.96 ,0.30) | 0.41 |
| | | L3 map volume | -0.18 (-0.45, 0.09) | 0.36 |
| | | L5 map volume | 0.06 (-0.24 ,0.37) | 0.69 |
| | Adjusted[1] | Log-transformed $N_{80}$ SEP area | -0.42 (-1.28, 0.45) | 0.44 |
| | | Log-transformed $N_{150}$ SEP area | -0.62 (-1.47, 0.23) | 0.29 |
| | | L3 map volume | -0.46 (-0.88, -0.04) | 0.13 |
| | | L5 map volume | -0.11 (-0.56, 0.35) | 0.63 |

[1] Adjusted for predisposing factors, blood biomarkers, psychological variables and sensitisation.
Statistically significant values are in bold font
B – unstandardized beta coefficient, CI – confidence interval, FDR – false discovery rate, SEP – somatosensory evoked potential

**Complete case analysis - Table 4.** Adjusted and unadjusted logistic regression models, including only complete cases, to test the effects of baseline sensorimotor activity on chronic pain and chronic disability at six-month follow-up.

| **Outcome** | **Model** | **Exposure** | **OR (95% CI)** | $\boldsymbol{P_{FDR}}$ | **E-Value** | **E CI** |
|---|---|---|---|---|---|---|
| Chronic pain [&] | Unadjusted | Log-transformed $N_{80}$ SEP area | 2.08 (1.52, 3.03) | **< 0.001** | 2.24 | 1.76 |
| | | Log-transformed $N_{150}$ SEP area | 2.04 (1.45, 2.94) | **< 0.001** | 2.21 | 1.70 |
| | | L3 map volume [n.b.] | 1.11 (0.98, 1.28) | 0.15 | 1.29 | 1.00 |
| | | L5 map volume | 0.99 (0.85, 1.15) | 0.91 | 1.08 | 1.00 |
| | Adjusted [1] | Log-transformed $N_{80}$ SEP area | 3.45 (1.30, 14.29) | 0.08 | 3.12 | 1.54 |
| | | Log-transformed $N_{150}$ SEP area | 7.14 (2.04, 100.00) | 0.07 | 4.79 | 2.21 |
| | | L3 map volume [n.b.] | 1.16 (0.93, 1.49) | 0.23 | 1.37 | 1.00 |
| | | L5 map volume | 1.12 (0.67, 2.00) | 0.61 | 1.31 | 1.00 |
| Chronic disability [#] | Unadjusted | Log-transformed $N_{80}$ SEP area | 1.37 (1.03, 1.85) | 0.07 | 1.62 | 1.14 |
| | | Log-transformed $N_{150}$ SEP area | 1.22 (0.92, 1.67) | 0.18 | 1.44 | 1.00 |
| | | L3 map volume [n.b.] | 1.25 (1.06, 1.52) | 0.04 | 1.48 | 1.21 |
| | | L5 map volume | 1.14 (0.98, 1.35) | 0.15 | 1.33 | 1.00 |
| | Adjusted [1] | Log-transformed $N_{80}$ SEP area | 2.00 (0.99, 5.00) | 0.22 | 2.18 | 1.00 |
| | | Log-transformed $N_{150}$ SEP area | 1.69 (0.93, 3.45) | 0.22 | 1.93 | 1.00 |
| | | L3 map volume [n.b.] | 1.16 (0.93, 1.56) | 0.23 | 1.37 | 1.00 |
| | | L5 map volume | 1.45 (0.93, 3.70) | 0.23 | 1.70 | 1.00 |

[&] Chronic pain in this logistic model was defined as NRS score ≥ 1 at 6-month follow up.
[#] Chronic disability in this logistic model was defined as RMDQ score ≥ 3 at 6-month follow up.
[1] Adjusted for predisposing factors, blood biomarkers, psychological variables and sensitisation.
[n.b.] L3 map volume logistic regression models were not adequately powered to detect a meaningful effect. Therefore, we controlled for age, DASS, TNF-α and local PPT, a single variable from each confounder category. Despite this, no meaningful effects were observed, therefore this is unlikely to impact study conclusions.
Statistically significant values are in bold font
CI – confidence interval; DASS – depression, anxiety, stress subscale; FDR – false discovery rate; OR – odds ratio; PPT – pressure pain threshold; SEP – somatosensory evoked potential; TNF – tumor necrosis factor.
